# Maternal Opioids Downregulate Adiponectin Receptor Signaling and Alter Growth in Offspring: Pilot Study

**DOI:** 10.64898/2026.01.08.26343734

**Authors:** Elizabeth Yen, Kiran Singh, Marissa Chow, Francesca Carasi-Schwartz, Mario Cordova, Tomoko Kaneko-Tarui, Emily Brew, Taysir Mahmoud, Pratik Reddy, Angie Mae Rodday, Jill Maron, Jonathan M. Davis, Perrie O’Tierney-Ginn

**Author notes:** **Corresponding author:** Elizabeth Yen, MD, 800 Washington Street, Box #44, Boston, MA 02111.

## Abstract

Opioid use disorder (OUD) has been linked to cardiometabolic diseases in adults through reductions in adiponectin—an adipocytokine with insulin-sensitizing effects. Opioid use during pregnancy dysregulates neonatal growth and may predispose to adult-onset diseases, but the impact of maternal OUD on neonatal adiponectin has not been studied. We hypothesize that maternal OUD also reduces adiponectin level in offspring (primary outcome) and alters growth (secondary outcome). To test our hypothesis, we conducted a prospective, observational pilot study and compared the expression of salivary adiponectin receptor 1/*ADIPOR1* and anthropometric and body composition (fat and fat-free mass) measurements between opioid-exposed and age-matched non-exposed neonates born at ≥34 weeks’ gestation. Data were stratified by exposure and sex using a Student’s t-test. Significance was set at p<0.05. A total of 67 neonates (35 opioid-exposed, 32 non-exposed neonates) were enrolled. Compared to healthy, non-exposed neonates, the expression of *ADIPOR1* was reduced in opioid-exposed neonates (0.27-fold, p<0.01), with the lowest expression in those requiring pharmacotherapy (0.048-fold, p<0.001). Despite the smaller anthropometric measurements in the exposed than non-exposed neonates (2915±625 grams vs. 3209±345 grams, p=0.02), opioid-exposed neonates had comparable adiposity to non-exposed neonates (8.60±4.52% vs. 8.53±4.00%, p=0.95). Less breast milk was used in the exposed than non-exposed group (25.7% vs. 71.9%, p<0.01). Maternal OUD may be associated with aberrant growth and excess adiposity in offspring through its effect on adiponectin signaling, predisposing these neonates to cardiometabolic risks.

## 1. INTRODUCTION

Neonates with *in-utero* opioid exposure are often born small and premature (1,2) and experience withdrawal signs called neonatal abstinence syndrome (NAS) or neonatal opioid withdrawal syndrome (NOWS), further affecting feeding and growth (3). Despite the initial growth deficits, these neonates reportedly experience a comparable growth trajectory as non-exposed neonates by four months (4). This “catch-up” growth resembles the growth pattern in neonates with intrauterine growth restriction (IUGR) or small for gestational age (SGA), placing them at risk for cardiovascular issues (5). Known as the “Barker hypothesis” or Developmental Origins of Health and Disease (DOHaD), undernutrition and adverse conditions during pregnancy are postulated to reprogram fetal development and metabolism and subsequently increase the risk of adult cardiac and metabolic disorders (6).

Adults with opioid use disorder (OUD) are at risk for insulin resistance and cardiometabolic diseases (7, 8), potentially through inhibition of adiponectin (a regulator in the insulin signaling pathway) (9). Adiponectin is an adipocyte-specific cytokine with insulin-sensitizing properties that exert anti-inflammatory, anti-obesity, and anti-atherogenic functions. Its action is mediated by adiponectin receptors (e.g., ADIPOR1, ADIPOR2) (10). Reduction in adiponectin is associated with insulin resistance, dyslipidemia, and atherosclerosis, and may account for the higher rates of metabolic syndrome and cardiovascular diseases in adults with OUD (11, 12). Further, opioid-related adiponectin suppression seemed to persist over time (13). Despite the known association between OUD and reduced adiponectin affecting cardiometabolic outcomes in adults, the effect of maternal OUD on offspring adiponectin has not yet been studied. In this pilot study, we hypothesize that maternal OUD is associated with decreased adiponectin level in offspring (primary outcome), affecting anthropometric and body composition measurements at birth (secondary outcome). Because receptor activation is vital for downstream cellular and physiological effects (14), we focused on adiponectin receptor rather than adiponectin itself. Given the sex-specific effects on gene expression levels in our study (15), we also explored whether sex modifies these relationships.

## 2. METHODS

Sample size calculation was not performed for this pilot study. We included neonates born at ≥34 weeks’ gestation in the Tufts Medicine Network between June 2022 and June 2024 whose families consented to saliva collection and/or body composition measurements as approved by the Tufts Medical Center Institutional Review Board. Opioid-exposed neonates were defined as those born to mothers with OUD by verbal report and/or positive maternal toxicology screening. The non-exposed cohort consisted of neonates without prenatal opioid exposure, with gestational age (GA) matched as close as possible between the two cohorts (within 1 week). Neonates with chromosomal or congenital anomalies were excluded.

### 2.1. Clinical and Demographic Data

Demographic data were extracted from electronic medical records (EMRs) to examine variables that may confound the effects of opioids on offspring adiponectin and growth measurements. Maternal factors collected include race, ethnicity, mode of delivery, tobacco use, group B streptococcus (GBS) status, hepatitis C infection status, type of opioid, and polysubstance use (defined as one or more additional psychotropic drugs in addition to opioids). Neonatal factors include sex, GA, Apgar scores at 1 and 5 minutes, anthropometric data (birth weight/BW, length/L, head circumference/HC, and corresponding percentiles), and SGA status (BW <10^th^ percentile) (16). Feeding types at discharge were categorized as exclusive breast milk use (BM), exclusive formula, or mixed. Opioid-exposed neonates were further categorized as those receiving pharmacotherapy (NAS Tx) and those not receiving pharmacotherapy (NAS No Tx) based on the Modified Finnegan Scoring System (17).

### 2.2. Salivary Gene Expression

All saliva samples were collected within 48 hours of birth. For opioid-exposed neonates, saliva collection occurred before the start of pharmacotherapy. Saliva was collected at least 1 hour before or after feeding to minimize milk contamination using previously described techniques (18). Briefly, neonates’ mouths were gently suctioned using a 1-milliliter (mL) insulin syringe (Becton, Dickinson and Company, Franklin Lakes, NJ) attached to the low-pressure wall suction for approximately 15 seconds (s). Saliva was immediately placed in Eppendorf tubes pre-filled with 250 microliter (μL) RNAprotect Saliva Reagent (Qiagen, Hilden, Germany) to minimize RNA degradation, then vortexed and stored at 4°C for at least 48 hours but no more than 28 days pending RNA extraction. RNA was extracted using the RNeasy Micro Kit (Qiagen) per the manufacturer’s instructions. To minimize DNA contamination, on-column DNase treatment was performed for each sample using RNase-free DNase I Set (Qiagen). Extracted RNA was stored at -80°C pending reverse transcription.

As receptor activation is important for physiological effects,^14^ we made *a priori* decision to measure adiponectin receptor expression. Further, the expression of adiponectin (*ADIPOQ*) was shown not to be reliably detected in neonatal saliva (19). Among the two adiponectin receptors, we targeted *ADIPOR1* based on its bioavailability and relative abundance in saliva (e.g., 103.5 number of transcripts per million (nTPM) for *ADIPOR1* vs. 21.6 nTPM for *ADIPOR2*) (20, 21). To properly normalize for varying starting total mRNA input across samples, we used two reference genes previously shown to remain stable across sex and post-menstrual age, i.e., glyceraldehyde-3-phosphate dehydrogenase (*GAPDH*) and tyrosine 3-mono-oxygenase/tryptophan 5-mono-oxygenase activation protein, zeta polypeptide (*YWHAZ*) (22). Threshold cycle (Ct) values for genes of interest and reference genes were detected using the QuantStudio^TM^ Flex Real-Time PCR (Applied Biosystems) machine. Normalized delta Ct (ΔCt) values were obtained by subtracting the geometrical mean Ct values of the housekeeping genes from the mean Ct values of the target gene (samples run in duplicate). The ΔΔCt values were derived by subtracting the ΔCt values of the exposed from the unexposed neonates. Relative fold change across exposure cohort was calculated using the ΔΔCt method (2^^(-meanΔΔCt)^) (23).

The predesigned forward and reverse primers for *ADIPOR1*, *GAPDH*, and *YWHAZ* were all purchased from MilliporeSigma (KiCqStart^®^ SYBR^®^ Green Primers). To determine the optimal working concentrations of the forward and reverse primers of each gene of interest along with the reference genes, we made a 100 micromolar (μM) stock solution for each primer according to the specifications determined by the company, followed by the preparation of working solutions in varying concentrations of 500 nanomolar (nM), 1 μM, 2 μM, 4 μM, and 8 μM for each forward and reverse primer. The desired/optimal combinations of primer concentrations for each gene are listed in **Table 1**.

**Table 1.**
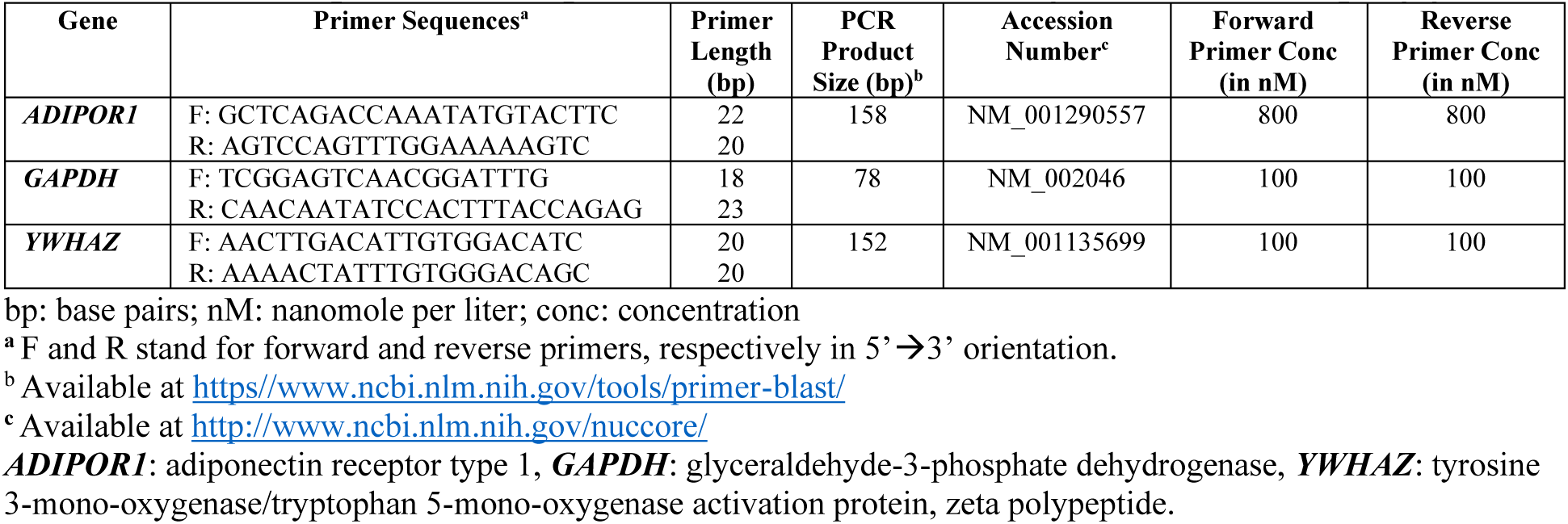
Primer sequences and optimal concentrations of target and housekeeping genes

Amplification mixture with a final volume of 10 μL per reaction was prepared by mixing 6 μL of the diluted cDNA with 10 μL of PowerUp^TM^ SYBR® Green Master Mix (Applied Biosystems, Carlsbad, CA), followed by 2 μL of each forward and reverse primer of each gene at the optimal concentrations as described above. Sample mix was plated on 96-well plate (Applied Biosystems) and run on the QuantStudio™ 7 Flex Real-Time PCR (Applied Biosystems) platform with the following thermal cycle profile: incubation at 50⁰C for 2 minutes (min) followed by one cycle of denaturation at 95⁰C for 10 min, then 40 cycles of 95⁰C for 15 s and 60⁰C for 60 s. At the end of the PCR, the temperature increased from 60⁰C to 95⁰C at a rate of 3⁰C per min. All samples were run in duplicate to achieve reproducibility. A commercial total control RNA (Applied Biosystems^TM^, Fisher Scientific) was used as a positive control and was included in all plates to assess plate-to-plate variability.

### 2.3. Body Composition Measurements

Body composition was measured using two non-invasive methods. The first was an air displacement plethysmography (ADP) method (PEA POD, COSMED, Italy), which is approved by the US Food and Drug Administration (FDA) for infants up to 6 months old and weighing up to 8 kilograms (kg). This system relies on a two-compartment model (body mass and body volume) to measure infant adiposity (fat- and fat-free mass) (24, 25) and is reliable in both term and preterm infants (24, 26). The ADP method has been validated against the gold standard four-compartment model and is highly accurate and reliable in determining infant body composition (26, 27), with no significant differences in the within- and between-day percent fat mass measurements, and is independent of the infant’s behavioral state (28, 29). Briefly, the PEA POD machine was calibrated before each use. Infants were placed naked on a scale inside the warmed chamber for 2 minutes. Weight was measured to the nearest 0.001 kg. Using Boyle’s Law, the machine calculated body composition measurements, including fat mass, fat-free mass, percent fat mass, percent fat-free mass, and body mass (30). Percent fat mass was converted to percentiles using the standardized body composition chart developed by Norris et al.^31^ Using this nomogram, percentiles of percent fat mass were normalized by gestational, postnatal, postconceptional age, and sex, accounting for fluid shift in the first few days of postnatal life (31, 32).

The second method for obtaining body composition involved using skinfold thickness and length measurements to estimate the amount of infant adiposity (33). The flank measurement was shown to be most reproducible and with the least variability compared to biceps, triceps, subscapular, and quadriceps measurements (34, 35). Using Harpenden calipers (Baty, United Kingdom), the flank skinfold was measured to the nearest 0.1 millimeter (mm) in the mid-axillary line just above the hip by carefully lifting the skin with thumb and index finger, avoiding the underlying tissue. Infant length was measured using a stadiometer (length board). Fat mass was calculated using the Catalano formula (0.54657 + [0.39055 * birth weight (kg)] – [0.03237 * birth length (cm)] + [0.0453 * flank skinfold (mm)]).^35^ The Catalano formula has been shown to accurately estimate neonatal fat mass when referenced against ADP (33, 35).

### 2.4. Statistical Analysis

Data analysis for this study was conducted using R Statistical Software (v4.4.0; R Core Team, 2024). Prism was used to create figures (v10.2.3; GraphPad, 2024). Statistical testing with a two-sided alpha of 0.05 was used for the primary and secondary outcomes, while other results were presented descriptively. Clinical and demographic data were described by opioid exposure. To determine the association between maternal opioid use and the expression of *ADIPOR1* as a marker of insulin sensitivity and growth measurements (anthropometric and body composition), we used linear regression with and without adjustment for maternal cigarette use. We also described growth measurements by opioid exposure. To evaluate the relationship between adiponectin signaling and growth, the salivary expression of *ADIPOR1* at birth was correlated with the anthropometric and body composition measurements using Pearson (parametric) or Spearman (non-parametric) correlation coefficients. For the correlation analyses, ΔCt values instead of the relative fold change (ΔΔCt) were used (36). Given the inverse relationship between ΔCt values and gene expression levels, a negative correlation represents a positive association, and vice versa (36). We also explored the differential effects of sex and NAS severity (defined as the need for pharmacotherapy) on the association between opioid exposure and primary and secondary outcomes. Given the small sample size, results were reported descriptively rather than including an interaction term between sex and opioid exposure.

To examine the congruence between anthropometric (BW, L, HC) and body composition measurements (fat mass, fat-free mass, skinfold thickness), we created a correlation matrix between these variables. Calculated fat mass using the skinfold measurement (Catalano formula) was also correlated with ADP-measured fat mass to determine how well these measurements correspond with one another.

## 3. RESULTS

Sixty-seven neonates were enrolled using the inclusion and exclusion criteria, comprising 35 opioid-exposed and 32 non-exposed neonates. A flow diagram of the subject enrollment is shown in **Figure 1**. Of the 67 enrolled neonates who consented to the saliva collection, 43 also consented to the body composition assessments.

**Figure 1.**
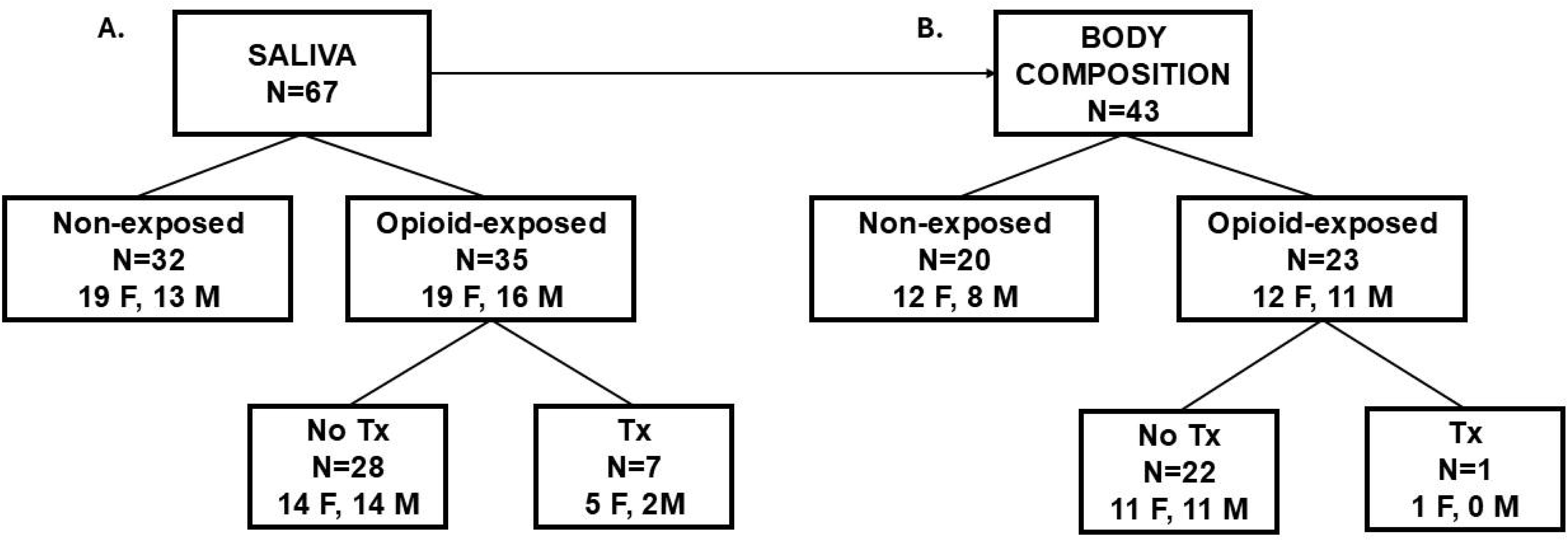
Flow Diagram of Subject Enrollment. Saliva samples were collected from 67 neonates at birth (A). Of these neonates, 43 underwent body composition measurements after birth (B). F=female, M=male, Tx=pharmacotherapy.

### 3.1. Clinical and Anthropometric Data

Demographic data of all 67 neonates in **Table 2** demonstrated that the exposed cohort was born smaller (BW, HC, and L), with a ten-fold incidence of SGA than non-exposed cohort. Significantly less breast milk and more formula or mixed feeding was used in the exposed cohort. Opioid-exposed neonates also stayed significantly longer (∼6 days) in the hospital, and about 20% required pharmacotherapy. The exposed cohort had more maternal hepatitis C status and tobacco use than the non-exposed cohort. For the type of MOUD, there was an equal distribution between buprenorphine and methadone use. More than two-thirds of women with OUD had polysubstance use. Stratification by sex shows that maternal opioid use was associated with greater reduction in the BW percentile and HC percentile in opioid-exposed females than males (**Table 3**).

**Table 2.** Demographic data of all subjects

|  | Non-Exposed (N=32) | Opioid-Exposed (N=35) | P<sup>1</sup> |
| --- | --- | --- | --- |
| Male sex | 13 (40.6) | 16 (45.7) | 0.640 |
| GA (weeks) | 38.3 (37.0, 39.1) | 37.6 (37.0, 39.1) | 0.209 |
| Race |  |  | 0.104 |
| White | 20 (62.5) | 24 (68.6) |  |
| Black | 0 (0.0) | 3 (8.6) |  |
| Other | 7 (21.9) | 2 (5.7) |  |
| Unknown | 5 (15.6) | 6 (17.1) |  |
| Ethnicity |  |  | 0.173 |
| Not Hispanic or Latino | 24 (75.0) | 25 (71.4) |  |
| Hispanic/Latino | 5 (15.6) | 2 (5.7) |  |
| Unknown | 3 (9.4) | 8 (22.9) |  |
| Cesarean section | 16 (50.0) | 16 (45.7) | 0.684 |
| 1-minute Apgar | 8.5 (8, 9) | 8 (7, 9) | **0.017** |
| 5-minute Apgar | 9 (9, 9) | 9 (9, 9) | 0.330 |
| BW (grams) | 3209 (345) | 2915.2 (625.1) | **0.022** |
| BW percentile | 59.5 (53.8, 66.3) | 46.0 (8.0, 63.0) | 0.074 |
| Length (cm) | 50.0 (3.6) | 47.9 (3.3) | **0.014** |
| Length percentile | 72.5 (45.3, 84.8) | 28.0 (9.0, 69.5) | **<0.001** |
| HC (cm) | 34.3 (2.7) | 32.9 (2.2) | **0.029** |
| HC percentile | 52.5 (32.5, 63.3) | 23.0 (10.5, 56.5) | 0.142 |
| Feeding |  |  | **<0.001** |
| BM | 23 (71.9) | 9 (25.7) |  |
| Formula | 4 (12.5) | 16 (45.7) |  |
| Mixed | 5 (15.6) | 10 (28.6) |  |
| SGA | 1 (3.1) | 12 (34.3) | **0.001** |
| LOS | 3 (2.6) | 8.9 (9.2) | **<0.001** |
| Pharmacotherapy for NAS | NA | 8 (22.9) | NA |
| Maternal GBS |  |  | **0.047** |
| Negative | 23 (71.9) | 22 (62.9) |  |
| Positive | 8 (25.0) | 5 (14.3) |  |
| Unknown/not done | 1 (3.1) | 8 (22.9) |  |
| Maternal hepatitis C |  |  | **<0.001** |
| Negative | 17 (53.1) | 14 (40.0) |  |
| Positive | 0 (0.0) | 15 (42.9) |  |
| Unknown | 15 (46.9) | 6 (17.1) |  |
| Maternal cigarette |  |  | **<0.001** |
| No | 25 (78.1) | 14 (40.0) |  |
| Yes | 2 (6.3) | 17 (48.6) |  |
| Unknown | 5 (15.6) | 4 (11.4) |  |
| Maternal medication |  |  | NA |
| Buprenorphine | NA | 15 (42.9) |  |
| Methadone |  | 16 (45.7) |  |
| Other |  | 4 (11.4) |  |
| Polysubstance | NA | 25 (71.4) | NA |

GA: gestational age, BW: birth weight, HC: head circumference, SGA: small for gestational age; LOS: length of stay; NA: not available/applicable, NAS: neonatal abstinence syndrome, GBS: group B streptococcus. Data are presented as mean (standard deviation) for normally distributed continuous measures, median (interquartile ranges) for non- normally distributed continuous measures, or N (%) for categorical measures. Bolded p values represent significance (p<0.05).

**Table 3.** Anthropometric measurements by sex and exposure

|  | **FEMALES (N=38)** |  | **MALES (N=27)** |  |
| --- | --- | --- | --- | --- |
|  | **Non-Exposed**<br>**(n=19)** | **Opioid-Exposed**<br>**(n=19)** | **Non-Exposed**<br>**(n=13)** | **Opioid-Exposed**<br>**(n=16)** |
| GA (wks) | 38.2 (1.4) | 37.8 (1.3) | 38.6 (1.7) | 37.5 (2.0) |
| BW (g) | 3,265.2 (355.5) | 2,865.7 (562.1) | 3,126.5 (324.8) | 2,973.9 (707.1) |
| BW percentile | 60.1 (23.5) | 40.7 (26.2) | 46.6 (26.3) | 43.8 (33.5) |
| HC (cm) | 33.8 (1.1) | 32.6 (1.8) | 33.2 (2.6) | 33.3 (2.5) |
| HC percentile | 51.6 (28.0) | 34.4 (28.2) | 46.8 (27.1) | 44.8 (33.7) |
| L (cm) | 50.6 (2.3) | 47.8 (2.9) | 50.3 (1.8) | 48.0 (3.9) |
| L percentile | 70.2 (26.0) | 43.1 (28.2) | 60.7 (27.8) | 42.5 (33.3) |

GA: gestational age, wks=weeks, BW: birth weight, g: grams, HC: head circumference, cm: centimeters, L: length, %: percentile. Data are presented as mean (standard deviation).

### 3.2. Salivary Gene Expression

**Table 4** demonstrated that maternal opioid use was associated with lower expression of *ADIPOR1* (0.27-fold, p<0.01), and this remained true after adjusting for maternal tobacco use (0.29-fold, p=0.02). Lower expression of *ADIPOR1* was observed in opioid-exposed males (0.18-fold compared to non-exposed males) and in opioid-exposed females (0.39-fold compared to non-exposed females) (**Figure 2A**, p<0.05). Within the exposed cohort, the expression of *ADIPOR1* was lower in females than males (0.45-fold, p=0.055). Stratification by the need for pharmacotherapy revealed a dose-response relationship, with the expression of *ADIPOR1* being lowest in the NAS Tx cohort (**Figure 2B**, p <0.05 for all comparisons).

**Figure 2.**
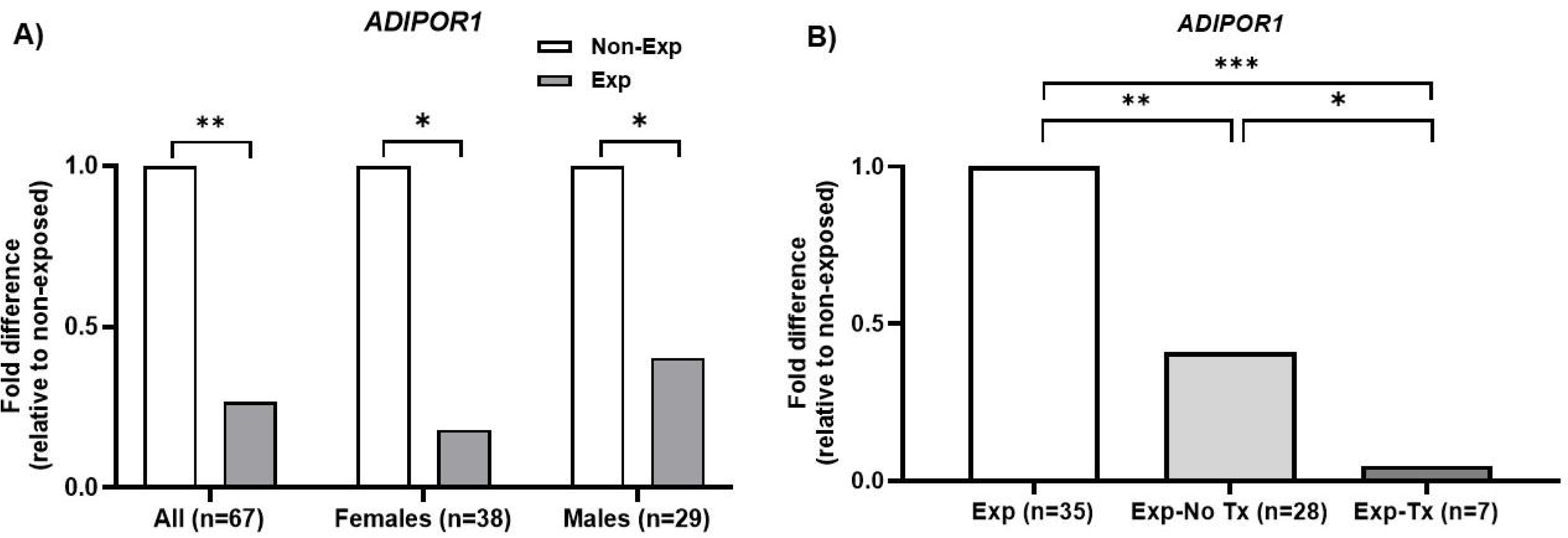
Evidence of Opioid-related Dysregulation of Insulin Sensitivity. A) The expression of *ADIPOR1* is significantly downregulated in opioid-exposed (NAS) than non-exposed neonates (No NAS), with similar effects seen in both males and females; B) Downregulation of *ADIPOR1* is most significant in the exposed neonates requiring pharmacotherapy (NAS Tx), suggesting that the impact of insulin sensitivity may be mediated by withdrawal severity. *ADIPOR1*=adiponectin receptor type 1; NAS=neonatal abstinence syndrome; Tx=treatment/pharmacotherapy. * p< 0.05, ** p< 0.01, *** p< 0.001

**Table 4.** Association between opioid exposure with *ADIPOR1* (N=67) and body composition (N=43) in unadjusted and adjusted linear regression models

|  | Beta (SE) | p-value | Fold expression <sup>a</sup> |
| --- | --- | --- | --- |
| ***ADIPOR1* ΔCt <sup>b</sup>** |  |  |  |
| Opioid exposure-unadjusted | 1.91 (0.63) | <0.01 | 0.27 |
| Opioid exposure-adjusted for smoking <sup>c</sup> | 1.78 (0.73) | 0.02 | 0.29 |
| **Percent Fat Mass** |  |  |  |
| Opioid exposure-unadjusted | 0.08 (1.31) | 0.95 |  |
| Opioid exposure-adjusted for smoking <sup>c</sup> | 0.47 (1.54) | 0.76 |  |
| **Percent Fat Mass Percentile <sup>d</sup>** |  |  |  |
| Opioid exposure-unadjusted | 4.20 (9.33) | 0.65 |  |
| Opioid exposure-adjusted for smoking <sup>c</sup> | 6.62 (10.98) | 0.55 |  |

<sup>a</sup> Fold expression calculated as $2^{-(\text{mean } ADIPOR1 \Delta Ct \text{ in opioid-exposed} - \text{mean } ADIPOR1 \Delta Ct \text{ in non-exposed})}$
<sup>b</sup> ΔCt is inversely proportional to gene expression level; a positive beta is a negative association.
<sup>c</sup> This model adjusts for smoking, and assumes those missing data on maternal cigarette use were non-smokers
<sup>d</sup> Normalized by GA, sex, postnatal age using method and charts by Norris et al.<sup>31</sup>

### 3.3. Body Composition Measurements

PEA POD and skinfold measurements were acquired in 20 non-exposed and 23 opioid-exposed. As shown in **Table 5**, body composition measurements were obtained significantly earlier in the non-exposed than opioid-exposed cohort (day 1 vs. 3). Opioid-exposed neonates had significantly lower body mass and lean mass, but comparable fat mass and skinfold measurements. To address the significant difference in the timing of measurements and potential effects of fluid shift in the first few days of life, we used Norris’ nomogram (31) to compare the percent fat mass percentiles. Linear regression models showed that opioid exposure was associated with non-significant increases in percent fat mass and percent fat mass percentiles (**Table 4)**. Descriptive analysis demonstrated the differential effect of sex on the relationship between opioid exposure and body composition, with reduced adiposity in opioid-exposed females, but increased adiposity in opioid-exposed males (**Table 6**).

**Table 5.** Body composition measurements after birth

| **ALL** | **Non-Exposed (N=20)** | **Opioid-Exposed (N=23)** | **P** |
| --- | --- | --- | --- |
| Timing (days of life) | 1.15 (0.67) | 2.96 (1.69) | **<0.01** |
| Body mass (kg) | 3.03 (0.36) | 2.69 (0.46) | **0.03** |
| Fat mass (kg) | 0.28 (0.15) | 0.25 (0.17) | 0.63 |
| Percent fat mass (%) | 8.53 (4.00) | 8.60 (4.52) | 0.95 |
| Percent fat mass percentile* | 35.77 (28.99) | 39.97 (31.78) | 0.66 |
| Fat-free/lean mass (kg) | 2.76 (0.28) | 2.44 (0.46) | **<0.01** |
| Percent fat-free/lean mass (%) | 91.47 (4.00) | 91.40 (4.52) | 0.95 |
| Skinfold (cm) | 3.74 (0.73) | 3.61 (1.07) | 0.66 |

kg: kilogram, cm: centimeter, \*normalized by GA, sex, postnatal age using Norris' nomogram.<sup>31</sup> Data were presented as mean (standard deviation). Bolded p values represent significance (p<0.05).

**Table 6.**
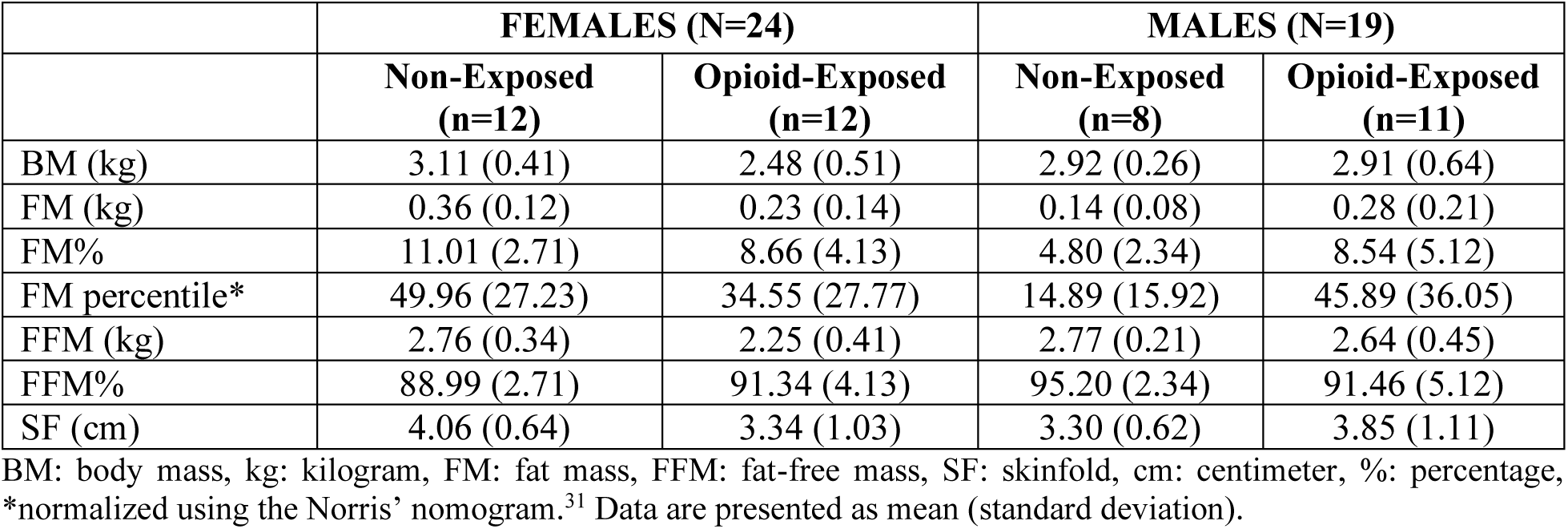
Body composition measurements by sex and exposure

|  | **FEMALES (N=24)** |  | **MALES (N=19)** |  |
| --- | --- | --- | --- | --- |
|  | **Non-Exposed**<br>**(n=12)** | **Opioid-Exposed**<br>**(n=12)** | **Non-Exposed**<br>**(n=8)** | **Opioid-Exposed**<br>**(n=11)** |
| BM (kg) | 3.11 (0.41) | 2.48 (0.51) | 2.92 (0.26) | 2.91 (0.64) |
| FM (kg) | 0.36 (0.12) | 0.23 (0.14) | 0.14 (0.08) | 0.28 (0.21) |
| FM% | 11.01 (2.71) | 8.66 (4.13) | 4.80 (2.34) | 8.54 (5.12) |
| FM percentile* | 49.96 (27.23) | 34.55 (27.77) | 14.89 (15.92) | 45.89 (36.05) |
| FFM (kg) | 2.76 (0.34) | 2.25 (0.41) | 2.77 (0.21) | 2.64 (0.45) |
| FFM% | 88.99 (2.71) | 91.34 (4.13) | 95.20 (2.34) | 91.46 (5.12) |
| SF (cm) | 4.06 (0.64) | 3.34 (1.03) | 3.30 (0.62) | 3.85 (1.11) |

BM: body mass, kg: kilogram, FM: fat mass, FFM: fat-free mass, SF: skinfold, cm: centimeter, %: percentage, \*normalized using the Norris' nomogram.<sup>31</sup> Data are presented as mean (standard deviation).

### 3.4. Association Between Salivary ADIPOR1 and Growth Parameters

Figure 3 demonstrates the correlations between *ADIPOR1* and BW percentile among non-exposed and opioid-exposed neonates. Lower expression of *ADIPOR1* (greater ΔCt values) correlated moderately with greater BW percentile in those with NAS (r = 0.28, panel **3B**), but this effect was mostly driven by severity of NAS (NAS Tx: r = 0.78, panel **3D**).

**Figure 3.**
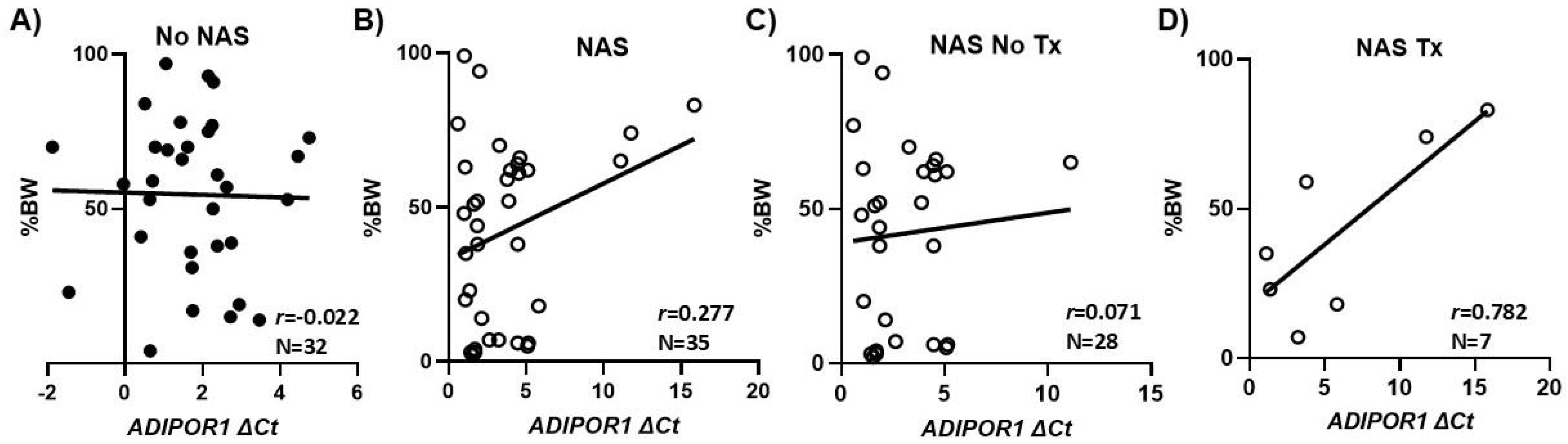
The Effect of Maternal Opioid Use on Insulin Sensitivity and Birth Weight. The expression of *ADIPOR1* trended with BW percentile in opioid-exposed neonates (NAS, panel B), with lower insulin sensitivity associated with greater birth weight at birth in those requiring pharmacotherapy (NAS Tx, panel D), suggesting a greater insulin signaling dysregulation in those with more severe withdrawal. NAS=neonatal abstinence syndrome, Tx=pharmacotherapy, BW%=birth weight percentile; *ADIPOR1*=adiponectin receptor type 1; ΔCt=delta threshold cycle, values are inversely proportional to the gene expression level.

### 3.4. Congruence of Anthropometric and Body Composition Measurements

Figure 4 shows the positive correlations between BW percentile and anthropometric (HC and L percentiles) and body composition (percent fat mass, skinfold) measurements in the non-exposed and opioid-exposed cohorts. Figure 5 demonstrates the heatmap of the rest of the correlation matrices between the anthropometric and body composition measurements in both non-exposed and opioid-exposed cohorts. Fat mass measured using the ADP methods correlated significantly with the skinfold-derived fat mass calculated using the Catalano formula (Figure 6).

**Figure 4.**
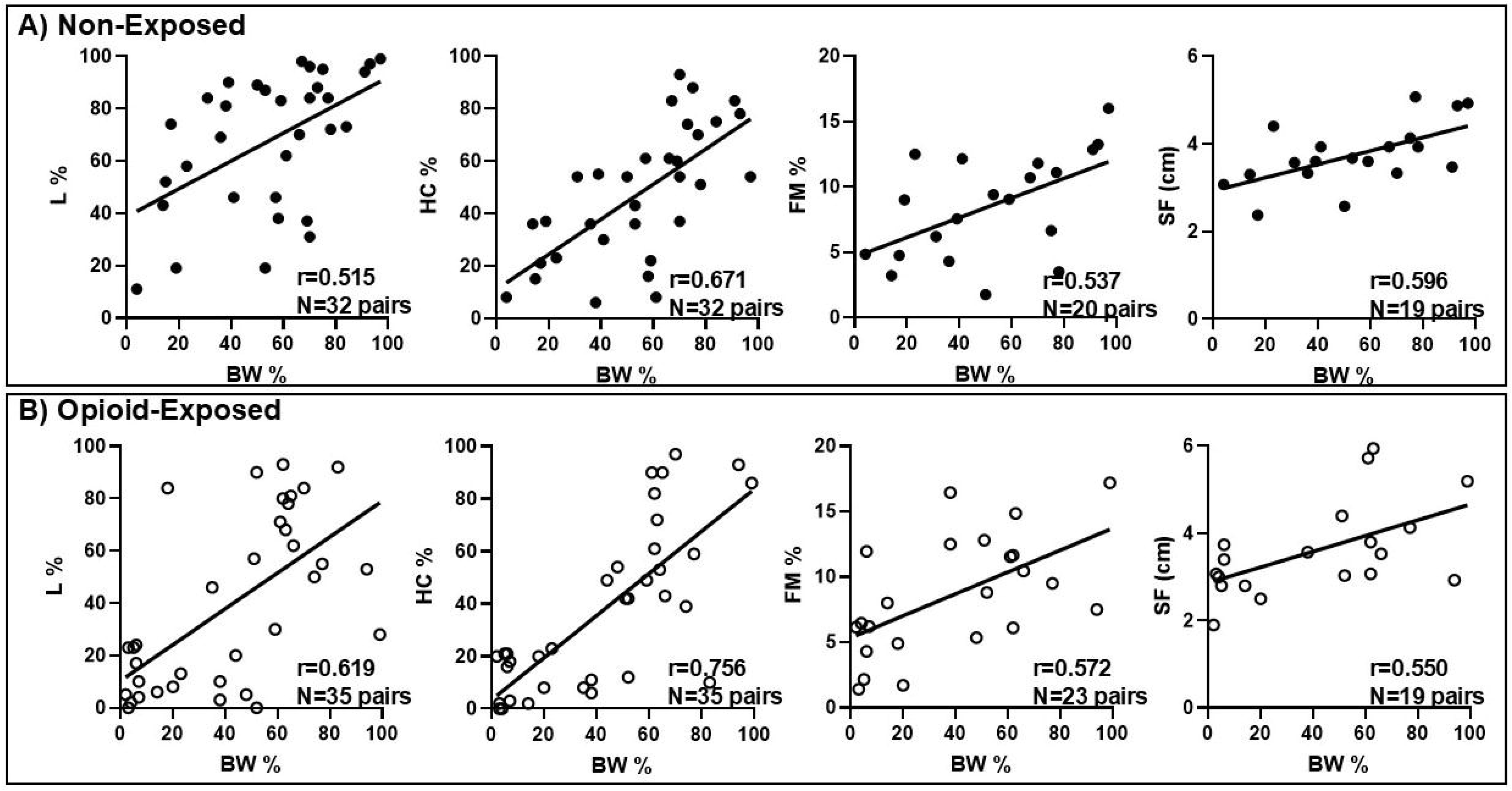
Anthropometric and Body Composition Measurements are Congruent in A) Neonates with no prenatal opioid exposure and B) Neonates with prenatal opioid exposure. L=length, HC=head circumference, FM=fat mass, SF=skinfold, %=percent, cm=centimeter.

**Figure 5.**
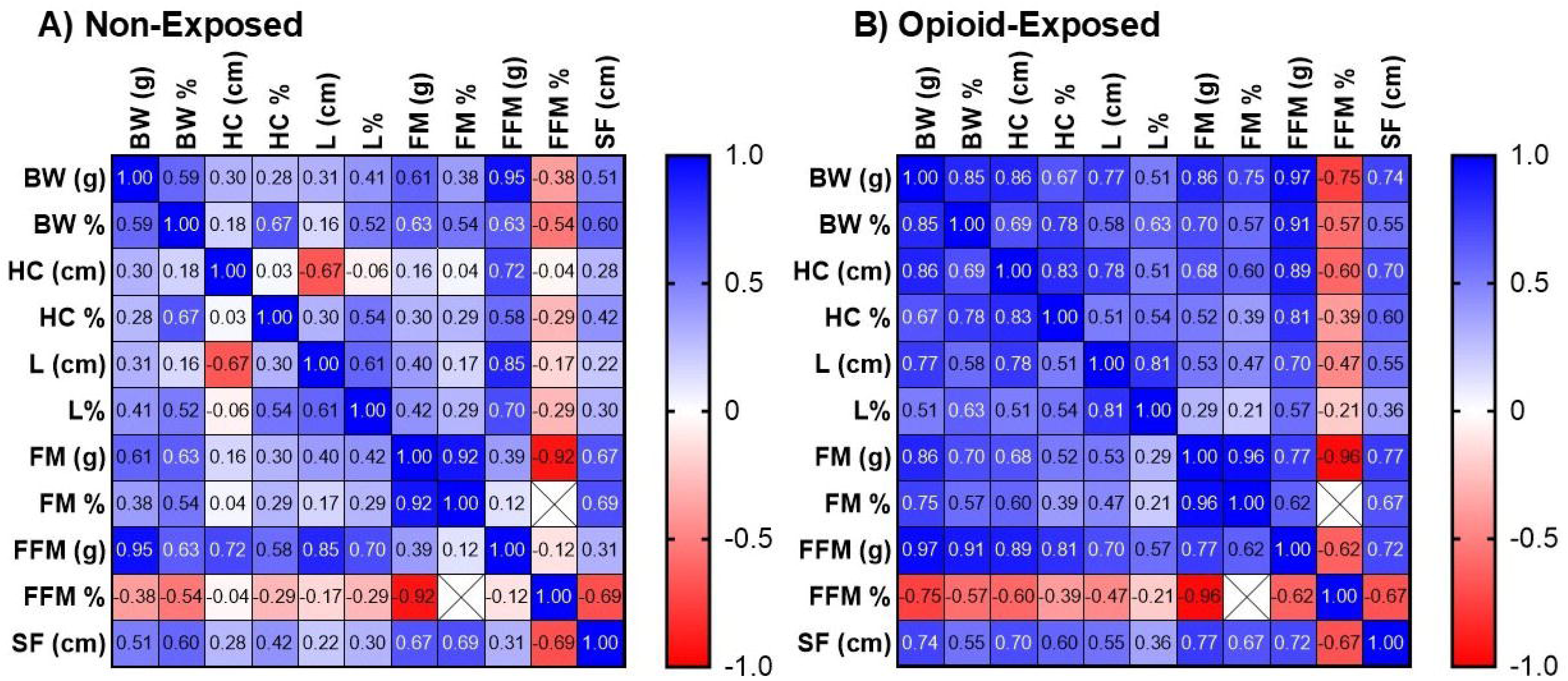
Correlation Matrices between Anthropometric and Body Composition Measurements. Heatmap diagrams show the correlations between anthropometric and body composition variables in non-exposed (A) and exposed (B) neonates. Blue indicates positive relationships, and red indicates negative relationships. Color intensity indicates the strength of relationships. Numerical values represent Pearson coefficients, with any values < 0.40 in either direction (negative or positive) being non-significant. BW=birth weight, HC=head circumference, L=length, FM=fat mass, FFM=fat-free mass, SF=skinfold, g=gram, cm=centimeter, %=percentile/percentage.

**Figure 6.**
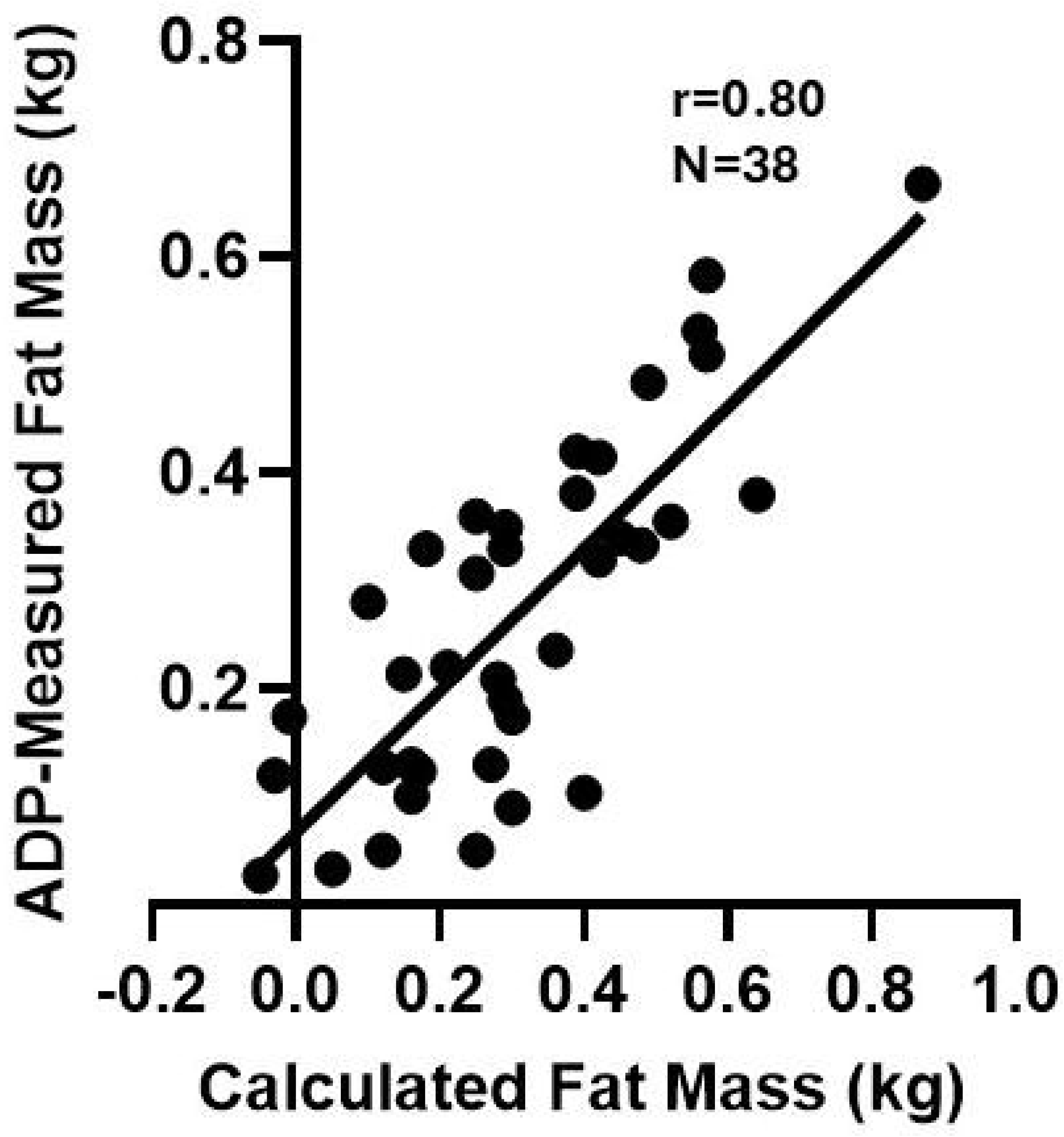
Congruence between Calculated and ADP-measured Fat Mass. Fat mass as measured by the ADP correlated significantly with skinfold-derived fat mass calculated using the Catalano formula. ADP: air displacement plethysmography; kg: kilogram.

## 4. DISCUSSION

Neonates exposed to opioids *in-utero* are often born smaller, but the mechanisms underlying this outcome are not well understood. Our observational pilot study demonstrates a potential molecular impact of maternal opioid use on neonatal adiponectin signaling underpinning growth aberrance in this population.

### 4.1. Primary Outcome: Lower Expression of ADIPOR1 in Opioid-exposed Neonates

Using the receptor activity to understand the physiological effects of maternal opioid use, our pilot study showed that opioid-exposed neonates had significantly lower salivary expression of *ADIPOR1*, suggesting that maternal opioid use may dysregulate adiponectin signaling in offspring. The action of adiponectin on its receptors holds an important role in insulin sensitivity, fetal growth, and development (10, 37). As a member of the adipokine family—adipocyte-derived cytokines and regulators of insulin function—adiponectin helps maintain metabolic health through anti-inflammatory effects, unlike the rest of the adipokines, which are pro-inflammatory (e.g., resistin, tumor necrosis factor alpha [TNFα], interleukin 6 [IL6], visfatin) (38, 39). Therefore, lower adiponectin levels have been linked to insulin resistance, metabolic syndrome, and type 2 diabetes mellitus (40, 41, 42).

The lower expression of *ADIPOR1* may also stem from the significantly higher proportion of SGA in the opioid-exposed cohort. Because neonates with SGA have less brown adipose tissue (BAT), and the adiponectin gene is highly expressed in BAT (43), it is plausible that the lower expression of *ADIPOR1* is mediated by the SGA status related to the prenatal opioid exposure. While we did not measure BAT in the current study, future studies can use imaging modalities (e.g., magnetic resonance imaging or spectroscopy) (44) to acquire BAT measurements. Importantly, lower adiponectin levels at birth have been inversely associated with changes in weight, length, and body mass index (BMI) in the first year and predict adiposity at 3 years of age (45, 46). Future studies should focus on the long-term growth trajectories of these neonates.

### 4.2. Secondary Outcome: Altered Anthropometric and Body Composition Measurements in Opioid-exposed Neonates

Our study demonstrated the correlation between body composition and anthropometric measurements. However, unlike anthropometrics, body composition measurements provide information on the composition and distribution of fat and fat-free/lean mass, which has been shown to predict future metabolic conditions (47, 48). Supporting the literature, opioid-exposed neonates in our study were born with a greater incidence of SGA and lower anthropometrics.^1^ SGA status has been negatively correlated with BMI and fat-free mass in childhood, with those born SGA undergoing body composition remodeling and faster adiposity accumulation when exposed to a nutrient-rich environment (49, 50). In addition to these anthropometric changes, the significantly lower fat-free/lean mass in opioid-exposed neonates may pose adverse risks. Lower lean mass in childhood has been associated with increased cardiometabolic risks in adulthood, as lean mass is an important determinant of normal growth, development, and glucose metabolism (51). Lower lean-to-fat mass ratio is also associated with the development of cardiovascular disease (52), suggesting that opioid-exposed neonates may be at risk for adverse cardiometabolic outcomes in adulthood.

Despite the significantly lower birth weight in opioid-exposed neonates, adiposity measurements (fat-mass, percent fat-mass, and percent fat-mass percentile) did not differ between groups. Adults with “normal weight obesity”—relatively greater body fat for the body weight—have an increased risk of developing metabolic syndrome, cardiovascular-related morbidities, or death (53). Our findings suggest that opioid-exposed neonates may have disproportionate fat- and lean-mass distribution at birth. Because weight deviation at birth may predispose to childhood adiposity and endocrine and metabolic disorders in adulthood (49, 53), opioid-exposed neonates may be at risk for adverse health outcomes. Further, the lower use of breast milk, which has a protective effect against excessive weight gain (54), may also exacerbate this long-term risk. Our pilot study also highlighted the potential use of body composition measurements as a valuable adjunct to anthropometric measurements by providing an estimate of adiposity.

### 4.3. The Link Between Lower ADIPOR1 and NAS Severity and Growth Measures

The dose-response relationship of the expression of *ADIPOR1* and NAS severity (Figure 2B) and the strong correlation between lower *ADIPOR1* and greater BW percentile in the NAS Tx cohort (Figure 3D) have not been previously reported. Maternal nutrition status may be a common pathway that modulates the relationships between adiponectin signaling, birth weight, and neonatal withdrawal severity. Studies have shown that both maternal malnutrition and overnutrition are associated with lower adiponectin levels and reduced insulin sensitivity in offspring (55, 56), as well as aberrant fetal growth through alterations in placental transport capacity and nutrient transfer (57). The current study did not examine maternal nutritional and placental factors, but our findings highlighted the need to understand the impact of opioids on the maternal-placental-fetal interactions and how these multifaceted interactions affect neonatal outcomes, both at the molecular level and in clinical presentations.

### 4.4. Sex-specific Effects of Maternal Opioid Use: Clinical and Molecular Evidence

Opioid-exposed females had lower anthropometric and adiposity measurements than non-exposed females. In contrast, opioid-exposed males were born with a comparable anthropometric size but greater adiposity than their non-exposed counterparts. Altogether, our results suggest that maternal opioid use is associated with reduced growth potential, primarily in females. Such a differential effect may result from a small sample size and random sampling error. Further validation is needed to confirm these findings. Given the increased cardiometabolic risks with either undernutrition or overnutrition, maternal opioid use may predispose either sex to aberrant cardiometabolic outcomes. Future research should examine the sex-specific effects of maternal opioid use on offspring growth and cardiometabolic outcomes in this high-risk population.

On the molecular level, our results showed that maternal opioid use lowered the expression of *ADIPOR1* in both males and females. Paralleling the differential growth measurements, our study also showed lower expression of *ADIPOR1* in opioid-exposed females than males, supporting prior animal and human studies.^58,59^ Interestingly, some studies have demonstrated the opposite effect, i.e., a greater effect in females than in males, or no effect across both sexes (52, 60, 61). Such variations may be due to differences in species (rodents vs. humans), age (neonatal vs. adult), sample size, and sample type (tissue, serum, vs. cord blood). Since lower adiponectin at birth is inversely associated with anthropometric changes in the first 12 months of age (43), opioid-exposed females may be at risk for greater weight changes in the future. Our findings highlighted sex-specific vulnerability to the adiponectin signaling and growth effects of maternal opioid use and the need for longitudinal monitoring for cardiometabolic outcomes.

The strengths of our study include the non-invasive nature and ease of salivary sample collection, making it feasible for serial analyses. With the molecular exchange between saliva and blood, saliva has become a promising biofluid that reflects systemic health (62). Given the early collection within 48 hours after birth, the changes in gene expression and insulin sensitivity are likely attributable to *in-utero* opioid exposure rather than postnatal effects. The use of body composition measurements, in addition to basic anthropometric data, provides valuable insights into the potential risk of poor cardiometabolic outcomes. The strong correlation between the ADP formula and the Catalano formula supports the utility of flank skinfold thickness to estimate neonatal adiposity in this population. Finally, our study also reveals molecular and clinical evidence of the sex-specific effects of maternal opioid use, further emphasizing the need to include sex as a biological variable in research pertinent to neonatal opioid withdrawal, among others (63).

Limitations of our study include a small sample size and a single-center design, which restrict the generalizability of our results. The one-time saliva and body composition acquisition also limits our understanding of the long-term effects of maternal opioid use. Not all families consented to body composition measurements, which may constrain the association between body composition and gene expression analyses. Finally, the current study lacks information on maternal nutritional and placental characteristics, which may influence adiponectin regulation in offspring and clinical presentations at birth. Therefore, our results should be interpreted cautiously.

### 4.5. Conclusion and Future Direction

This pilot study elucidates the molecular impact of maternal opioid use on adiponectin signaling and infant growth. Since adiponectin has insulin-sensitizing properties and insulin is a crucial intrauterine growth factor (9), lower expression of *ADIPOR1* and disproportionate size and adiposity at birth may predispose these opioid-exposed neonates to an unhealthy growth trajectory and future cardiometabolic issues. Therefore, our study provides objective evidence on how maternal opioid use may program developmental trajectory and health across the lifespan. Future studies should validate the current findings using a larger sample and over a longer period. Serial molecular and body composition data will improve our understanding of the mechanisms by which opioid-related adiponectin dysregulation affects cardiometabolic health. Mechanistic research in this field is urgently needed to improve and prevent public health issues that may result from the opioid epidemic, such as obesity, metabolic syndrome, and cardiovascular diseases.

## Data Availability

All data produced in the present study are available upon reasonable request to the authors.

## Abbreviation List

ADP: air displacement plethysmography
*ADIPOR1*: adiponectin receptor 1
BMI: body mass index
BW: birth weight Ct: threshold cycle
GA: gestational age
HC: head circumference
L: length
NAS: neonatal abstinence syndrome
NAS Tx: NAS requiring pharmacotherapy
OUD: opioid use disorder
SGA: small for gestational age

## Acknowledgment

We would like to thank all the families for participating in this study and the clinical leaders and staff at Tufts Medicine (Tufts Medical Center and Lowell General Hospital) for their support.

## Author Contributions

Conceptualization—EY, JM, JMD, PO-G Investigation—EY

Software—EY, AMR, PR

Data curation—KS, MCh, FC-S, MCo, EB, TM Methodology—EY, TK-T, AMR Supervision—EY, MCo, TK-T, AMR, PO-G

Formal analysis—EY, FC-S, MCh, TK-T, PR, AMR, PO-G Project administration—KS, TM

Funding acquisition—EY Resources—MCo, JM, JMD, PO-G Original drafting—EY

Review, editing, and final approval of the version to be published—EY, KS, FC-S, MCh, TK-T, MCo, EB, TM, PR, AMR, JM, JMD, PO-G

## Consent Statement

Informed consents were obtained from all subjects enrolled in this study as approved by the Tufts Institutional Review Board.

## Data Availability

Materials described in the manuscript, including all relevant raw data, are available for research and non-commercial purposes upon reasonable request, without breaching participant confidentiality.

## Funding Acknowledgement

National Institute on Drug Abuse (5K23 DA056847) (EY), Tufts Institute for Global Obesity Research (EY), and National Center for Advancing Translational Sciences (UM1TR004398) (Tufts CTSI). The funder did not play any role in the 1) study design, 2) the collection, analysis, and interpretation of the data; 3) the writing of the report; and 4) the decision to submit the manuscript for publication.

